# Independent and interactive effects of viral species on risk for lower respiratory tract illnesses in early life

**DOI:** 10.1101/2025.02.25.25322678

**Authors:** Camille M. Moore, Elizabeth A. Secor, Ana Fairbanks-Mahnke, Jamie L. Everman, Jennifer R. Elhawary, Jonathan I. Witonsky, Elmar Pruesse, Chih-Hao Chang, Maria G. Contreras, Celeste Eng, Keyshla Canales, Tsunami Rosado, Donglei Hu, Scott Huntsman, Nathan D. Jackson, Yingchun Li, Natalie Lopez, Annette Medina Valentin, Vivian Medina, Chris Angely Montanez-Lopez, Andrew Morin, Natalie A. Nieves, Sam S. Oh, Richeliz Alfonso Otero, Raymarie Colon, Leysha Rodriguez, Satria P. Sajuthi, Sandra Salazar, Gonzalo Serrano, Emily Vazquez Morales, Gabriela Vazquez, Nicole Vazquez Morales, Blake J. M. Williams, Priscilla Zhang, Dean Sheppard, Jose R. Rodriguez Santana, Max A. Seibold

## Abstract

**Importance:** All children experience upper respiratory tract illnesses (URI) caused by viral infections. However, some of these illnesses progress to the lower airways. Although studies have found infection with certain viral species are more likely to trigger lower respiratory illnesses (LRIs), a comprehensive analysis of viruses underlying early-life LRIs is lacking.

**Objective:** Determine the incidence of URIs, mild and severe LRIs (mLRI, sLRI) during the first 2 years of life and the association between viral respiratory pathogens and odds of LRIs versus URIs in Puerto Rican children, a population at high risk for respiratory disease.

**Design, Setting, and Participants:** Healthy mother-infant pairs were enrolled in the Puerto Rican Infant Metagenomic and Epidemiologic Study of Respiratory Outcomes birth cohort, in Caguas, Puerto Rico. Infants (n=2,061) were surveilled for respiratory illnesses during the first two years of life (March 2020 to April 2024). Nasal swabs from a subset of 1,363 illnesses from 774 participants were screened for 21 pathogens.

**Exposures:** Infection with respiratory pathogens.

**Main Outcomes and Measures:** URI, mLRI, and sLRI in the first two years of life.

**Results:** RSV infections occurred in 23% of sLRIs and were associated with dramatically increased odds of sLRI vs URI (OR=9.28; 95% CI, 5.43-15.85). Metapneumovirus, parainfluenza, and non-SARS-CoV-2 coronavirus infections also increased odds of sLRIs. SARS-CoV-2 was associated with lower risk of sLRIs vs. URIs (OR=0.33; 95% CI, 0.16-0.68). Though rhinovirus (43%) and bocavirus (16.1%) were commonly detected in sLRIs, neither was associated with increased sLRI risk. Infection with multiple viral species (i.e. co-infection) occurred in one-third of sLRIs and was associated with 2.92-fold greater odds of sLRI (95% CI, 2.05-4.16) compared to single viral species infections. Rhinovirus-bocavirus was the most common co-infection (32.4%), and interaction between these viral infections was associated with increased sLRI risk (OR=2.21; 95% CI, 1.20-4.09) relative to illnesses that were negative for rhinovirus and bocavirus.

**Conclusions and Relevance:** A diversity of viral pathogens drive early-life sLRIs. Some viral pathogens (e.g. RSV and metapneumovirus) have intrinsic propensity to cause sLRIs, whereas many sLRIs are caused by viruses whose lower airway pathogenicity is dependent on other factors, including co-infection.

**Key Points:** *Question:* How do common respiratory viruses differ in their prevalence and risk of causing severe lower respiratory illnesses (LRIs) during early childhood?

*Findings:* RSV, metapneumovirus, and parainfluenza are independent risk factors for early childhood severe LRIs. While rhinovirus and bocavirus infections alone do not increase the risk of severe LRIs, these two viruses significantly elevate risk when they occur as co-infections.

*Meaning:* Our findings highlight significant variability in viruses that drive severe early-life LRIs. Some viral species appear to inherently predispose individuals to lower airway disease, while for others, the development of disease likely depends on co-infections and/or host susceptibility.

## INTRODUCTION

Viral respiratory illnesses are common in early life. While many infections result in mild upper respiratory tract illnesses (URIs), others progress to lower respiratory illnesses (LRIs), characterized by cough, wheezing, and breathing difficulties, sometimes requiring corticosteroid treatment or hospitalization. LRIs can have lasting health impacts, including increased risk for asthma^1–6^, allergic sensitization^7^, recurrent respiratory infections^8^, and lung function deficits later in childhood.^9^ Therefore, identifying risk factors for the progression of viral infections to LRIs is critically important.

Host factors like prematurity and genetics influence LRI risk, but viral species are also key contributors.^10–12^ For example, respiratory syncytial virus (RSV) is strongly linked to infant bronchiolitis, while human rhinovirus (HRV)-A and HRV-C are common in wheezing illnesses.^10–16^ Few studies have investigated the relative effects of RSV vs. HRV infections. Advances in molecular detection, such as multiplexed PCR assays, have identified less frequent viruses, such as enterovirus, parainfluenza and metapneumovirus, as potential LRI risk factors. Broader screening has also shown that infections involving multiple viruses are common, though their impact is unclear, with studies showing conflicting outcomes ranging from increased ICU admissions to reduced severity.^17, 18^ Finally, the emergence of SARS-CoV-2 presents a new viral pathogen that could drive early-life LRIs, although population-based studies have indicated that SARS-CoV-2 is more likely to cause asymptomatic than symptomatic infections in young children.^19^ A large, early-life surveillance study with comprehensive viral screening is needed to clarify the LRI risks and prevalence of all common viral species.

The Puerto Rican Infant Metagenomic and Epidemiologic Study of Respiratory Outcomes (PRIMERO) was established to address these knowledge gaps in a population experiencing more severe early-life respiratory illnesses and bearing an exceptionally high burden of asthma.^20, 21^ This longitudinal cohort of 2,061 newborns monitored respiratory illnesses (URIs and LRIs) during the first two years of life. We report the incidence of URIs, mild LRIs (mLRIs), and severe LRIs (sLRIs) requiring hospitalization or systemic steroid treatment. Nasal swabs from 430 URIs, 319 mLRIs, and 577 sLRIs were screened for respiratory viruses to evaluate associations with mLRI and sLRI risk. Given that surveillance began during the COVID-19 pandemic, we also examined the impact of mitigation measures on LRI incidence and assessed the role of SARS-CoV-2 in early-life LRI risk.

## METHODS

### Study design

PRIMERO is a prospective birth cohort study of early-life respiratory illnesses and has been previously described in full.^22^ Briefly, healthy mother-infant pairs were enrolled from the Hospital Interamericano de Medicina Avanzada-San Pablo in Caguas, Puerto Rico. Respiratory illness surveillance was performed for the first 2 years of life via weekly SMS text and email messages. Mothers reporting that their child was ill were contacted by phone for LRI screening based on the presence of symptoms, including a cough interfering with daily activities, wheezing/whistling in the chest, fast breathing or gasping for air, and/or sleep disturbed by cough, wheeze, or difficulty breathing. LRIs were considered severe if they required hospitalization or treatment with systemic corticosteroids. Illnesses reporting only a runny/plugged nose, sneezing, and/or a mild cough were classified as URIs.

All LRIs, as well an infant’s first URI each year of life, were invited for an in-person assessment, during which nasal swabs were collected for screening with a multiplex PCR assay for 21 common respiratory viruses.

The PRIMERO protocol and informed consent documents were approved by the UCSF Institutional Review Board (IRB) and by a National Institutes of Health (NIH)-appointed independent Observational Study Monitoring Board (OSMB).

### Statistical analysis

We tested for association between viral species detection and odds of sLRI vs URI, mLRI vs. URI, and sLRI vs mLRI using generalized estimating equation logistic regression models, adjusting for age, sex, race, and time from illness onset to nasal swab. The model included indicator variables for the following viral detections: adenoviruses, coronaviruses other than SARS-CoV-2, bocaviruses, metapneumoviruses, rhinoviruses/enteroviruses, parainfluenzas, respiratory syncytial viruses, and SARS-CoV-2. We adjusted for statistical testing of the 8 virus types using a Benjamini-Hochberg correction to control the false discovery rate (FDR). For co-infections detected in >3% of illnesses, we tested for effect modification between pairs of viruses involved in co-infections by adding interaction terms to the model described above. Similar models compared the odds of mLRI and sLRI vs. URI between illnesses with no, single, and multiple viral detections.

Additional details are available in the eMethods.

## RESULTS

### Respiratory illness surveillance

2,061 children were surveilled for respiratory illnesses between March 3, 2020 and April 29, 2024, with a median follow-up of 24 months (IQR: 19 to 24, Table 1). 6,051 respiratory illnesses were reported and documented, 61.5% (n=3,721) and 38.4% (n=2,325) of which were URIs and LRIs, respectively (Figure 1A; 5 illnesses could not be classified). Of the 2,185 LRIs with severity data, 43.9% (n=960) of LRIs required hospitalization or systemic corticosteroid treatment and were classified as severe. Since illness surveillance spanned the period from the onset of the COVID-19 pandemic—when public health measures were in place to mitigate viral transmission—to the easing of these restrictions, we compared respiratory illness rates between children born during the first year of the pandemic (n=535, March 2020 to February 2021) with children born after restrictions were eased and schools reopened (n=1,526, March 2021 to April 2024). Using Kaplan Meier analyses, we estimated the probability of experiencing one or more URIs or LRIs during the first 2 years of life was 67.8% and 44.0%, respectively, for children born during the first year of the pandemic (Figure 1B). In contrast, the probabilities of these illnesses were significantly higher (URI: 72.1%, LRI: 56.2%) in children born after restrictions were eased. Among children with complete 2-year surveillance, 31.0% (n=154/497) and 21.9% (n=109/497) of children born during the first year of the pandemic reported one or more mLRIs and sLRIs, respectively; while 42.2% (n=322/763) and 35.4% (n=270/763) of those born after restriction easing reported mLRIs and sLRIs (eFigure 1).

**Figure 1.**
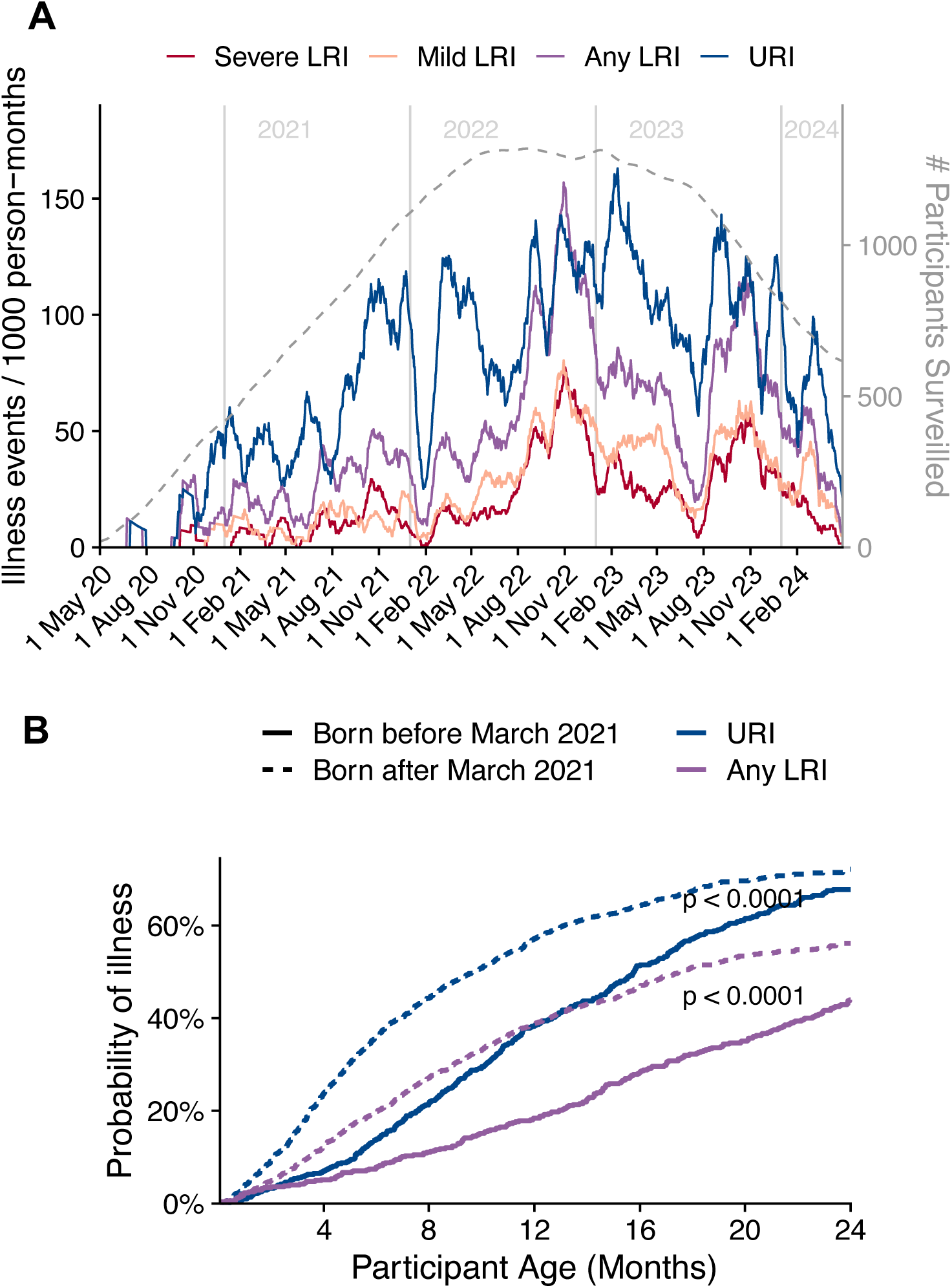
Respiratory illness surveillance from March 3, 2020 to April 29, 2024. A) 30 day rolling incidence of upper respiratory illnesses (blue), mild lower respiratory illnesses (orange), and severe lower respiratory illnesses (red) in the PRIMERO cohort from March 3, 2020 to April 29, 2024. The number of illness events per 1,000 person months was calculated as the sum of the number of ill ness events divided by the average number of participants surveilled in a window 15 days before to 15 days after the date times 1,000. The grey dashed line indicates the number of participants under active surveillance. The red shaded area indicates the first year of the COVID-19 pandemic (March 3, 2020 to March 1, 2021). B) Kaplan-Meier estimates and 95% confidence intervals for the probability of experiencing one or more URIs (blue) and LRIs (purple) from birth to 24 months of age for children born during the first year of the COVID-19 pandemic (solid lines) and children born after March 1, 2021 (dashed lines). P-values comparing curves between those born before and after March 1, 2021 are from the log-rank test.

**Table 1.** Characteristics of participants enrolled in the PRIMERO cohort stratified by whether a respiratory illness was reported and for participants reporting one or more illnesses, whether the participant ever had an in-person illness assessment. N (%) or median [interquartile range].

|  | Reported Illness |  | In Person Illness Assessment with Successful Pathogen Testing |  |
| --- | --- | --- | --- | --- |
|  | No (N=478) | Yes (N=1,577) | No (N=803) | Yes (N=774) |
| **Male Sex** | 1029 (49.9) | 792 (50.2) | 380 (47.3) | 412 (53.2) |
| **Race:** |  |  |  |  |
| **Black** | 106 (22.2) | 345 (21.9) | 167 (20.8) | 178 (23.0) |
| **Other/Mixed** | 182 (38.2) | 620 (39.3) | 314 (39.1) | 306 (39.6) |
| **White** | 189 (39.6) | 611 (38.8) | 322 (40.1) | 289 (37.4) |
| **Born During First Year of Pandemic** | 134 (28.0) | 397 (25.2) | 165 (20.5) | 232 (30.0) |
| **Follow Up (months)** | 24<br>[16.4, 24.0] | 24<br>[20.0, 24.0] | 24<br>[17.1, 24.0] | 24<br>[23.7, 24.0] |
| **Respiratory Illness Reported** | 0 (0.0) | 1577 (100.0) | 803 (100.0) | 774 (100.0) |
| **URI Reported** | 0 (0.0) | 1396 (88.5) | 696 (86.7) | 700 (90.4) |
| **LRI Reported** | 0 (0.0) | 1020 (64.7) | 341 (42.5) | 679 (87.7) |
| **Age at First Respiratory Infection (months)** |  | 5<br>[3.0, 10.0] | 6<br>[3.00, 12.00] | 4<br>[2.00, 8.00] |
| **Age at First Reported URI** |  | 6<br>[3.0, 11.0] | 7<br>[3.00, 12.00] | 6<br>[3.00, 10.00] |
| **Age at First Reported LRI** |  | 8<br>[4.0, 14.0] | 8<br>[4.00, 14.00] | 8<br>[4.00, 13.00] |
| **Number of URIs Reported** |  |  |  |  |
| 0 | 478 (100) | 181 (11.5) | 107 (13.3) | 74 (9.6) |
| 1 | 0 (0.0) | 502 (31.8) | 350 (43.6) | 152 (19.6) |
| 2 | 0 (0.0) | 324 (20.5) | 166 (20.7) | 158 (20.4) |
| 3 | 0 (0.0) | 206 (13.1) | 85 (10.6) | 121 (15.6) |
| 4 | 0 (0.0) | 154 (9.8) | 58 (7.2) | 96 (12.4) |
| 5 or more | 0 (0.0) | 210 (13.3) | 37 (4.6) | 173 (22.4) |
| **Number of LRIs Reported** |  |  |  |  |
| 0 | 478 (100) | 557 (35.3) | 462 (57.5) | 95 (12.3) |
| 1 | 0 (0.0) | 453 (28.7) | 241 (30.0) | 212 (27.4) |
| 2 | 0 (0.0) | 256 (16.2) | 75 (9.3) | 181 (23.4) |
| 3 | 0 (0.0) | 123 (7.8) | 14 (1.7) | 109 (14.1) |
| 4 | 0 (0.0) | 73 (4.6) | 8 (1.0) | 65 (8.4) |
| 5 or more | 0 (0.0) | 115 (7.3) | 3 (0.4) | 112 (14.5) |

### Viral infection rates and seasonality

In-person assessments and nasal swab collection were performed on 1,587 RIs (26.2%, eFigure 2). Nasal swabs were screened with a multiplex PCR assay for 21 common respiratory viruses. Of the 1,360 successfully assayed illnesses with complete covariate data, 74.1% tested positive for one or more viruses. Only human rhinovirus/enterovirus (HRV/EV) and SARS-CoV-2 were detected during the first year of the pandemic (Figure 2B). Shortly after restrictions were eased, all common respiratory viruses were detected, including respiratory syncytial virus (RSV), parainfluenza (PIV), influenza (INF), common coronaviruses (CoV), adenoviruses (AV), human metapneumovirus (HMPV), and human bocavirus (HBoV). Examination of circulation patterns (Figure 2B; eTable 2) revealed significant seasonal fluctuations in RSV, HBoV, and CoV. In particular, RSV and HBoV circulation were increased in the fall and winter. CoV circulation was higher in the winter and spring.

**Figure 2.**
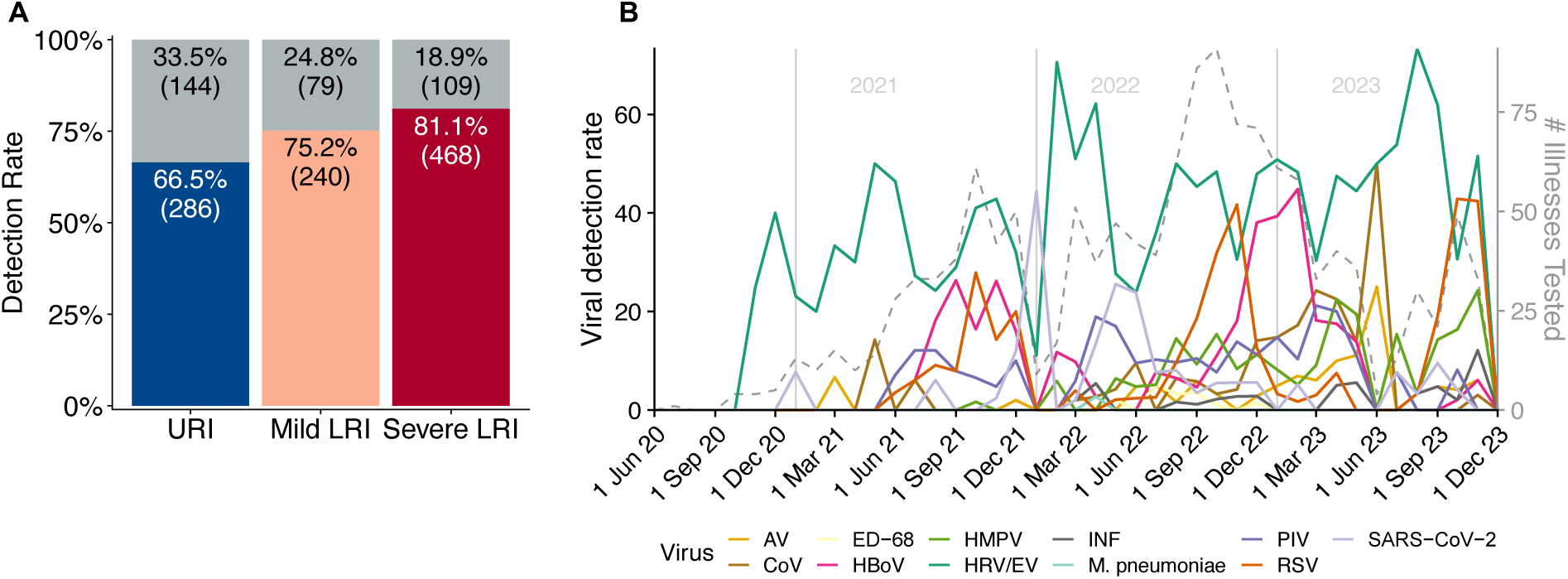
Viral detections in early life respiratory illnesses. A) Rates of viral detection among URIs (blue), mild LRIs (orange) and severe LRIs (red). B) Detection of viral species per month among illnesses assessed in-person. The viral detection rate is calculated as the number of viral positives divided by the number of illnesses successfully tested for all viruses during that month.

Among the 1,360 successfully screened respiratory illness were 430 URIs, 319 mLRIs, 577 sLRIs (34 LRIs with missing severity data, eTable 1). SLRIs were more likely to test positive for a virus than URIs (81.1% of sLRIs vs. 66.5% of URIs; adjusted OR (aOR), 1.63; 95% CI, 1.20 to 2.21; P=0.0019; Figure 2A), but not mLRIs (75.2% of mLRIs; aOR, 1.41; 95% CI, 1.00 to 2.01; P=0.053). Viral detection did not significantly differ between mLRIs and URIs (aOR, 1.13; 95% CI, 0.80 to 1.60; P=0.49).

### Viral species infections and LRI risk

#### HRV vs. RSV infections

HRV/EV was the most common viral species detected among both mLRIs and sLRIs (43.3% and 43.0%, respectively, Table 2). However, HRV/EV detections were not significantly associated with increased LRI odds relative to URIs (mLRI vs URI: aOR, 1.02, FDR=0.89; sLRI vs URI: aOR, 1.15; FDR=0.38), as HRV/EV infections also occurred in 40.9% of URIs. HRV subtyping was successfully performed on 84% (N=481) of HRV/EV positive illnesses, finding 50.3%, 3.7%, and 44.5% of illnesses were positive for HRV-A, HRV-B, and HRV-C, respectively. Neither HRV-A, HRV-B, nor HRV-C infections were individually associated with increased risk of LRIs (eFigure 3).

**Figure 3.**
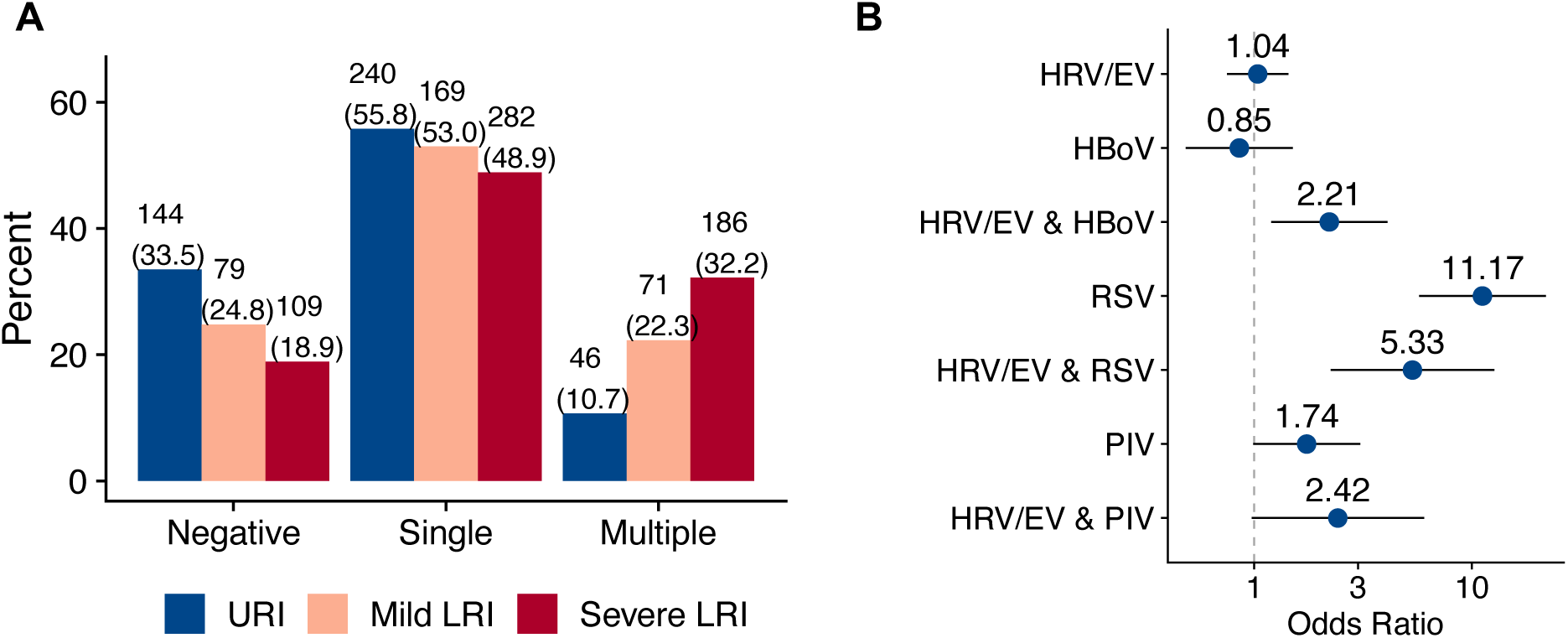
Association of co-infection with illness severity. A) Frequency of negative, single and multiple viral detections in URIs (blue), mild LRIs (orange) and severe LRIs (red). Each bar is annotated with N (%). B) Forest plot of adjusted odds ratios comparing the odds of severe LRI vs URI between illnesses that test positive and negative for specific viral infections and co-infections. Blue dots represent the estimated aOR and whiskers indicate the 95% confidence interval.

**Table 2.** Viral detections among URIs (N=432), mLRIs (N=286), and sLRIs (N=563) and odds ratios comparing the odds of mild or severe LRI between illnesses that test positive and negative for each viral species, adjusting for age at illness onset, sex, race, and time from illness onset to nasal swab. FDR: Benjamini-Hochberg adjusted p-value.

|  | URI<br>N (%) | mLRI<br>N (%) | sLRI<br>N (%) | mLRI vs URI |  | sLRI vs URI |  | sLRI vs mLRI |  |
| --- | --- | --- | --- | --- | --- | --- | --- | --- | --- |
|  |  |  |  | aOR<br>(95% CI) | FDR | aOR<br>(95% CI) | FDR | aOR<br>(95% CI) | FDR |
| **Adenoviruses** | 10 (2.3) | 13 (4.1) | 22 (3.8) | 1.53<br>(0.63, 3.68) | 0.556 | 1.37<br>(0.64, 2.96) | 0.417 | 0.84<br>(0.39, 1.79) | 0.684 |
| **Coronaviruses**<br>**(non-SARS-CoV-2)** | 14 (3.3) | 23 (6.9) | 45 (7.8) | 1.81<br>(0.83, 3.96) | 0.369 | 2.21<br>(1.22, 4.01) | **0.015** | 1.22<br>(0.7, 2.1) | 0.684 |
| CoV-229E | 1 (0.2) | 5 (1.6) | 5 (0.9) |  |  |  |  |  |  |
| CoV-HKU1 | 2 (0.5) | 3 (0.9) | 6 (1.0) |  |  |  |  |  |  |
| CoV-NL63 | 0 (0.0) | 4 (1.3) | 17 (2.9) |  |  |  |  |  |  |
| CoV-OC43 | 11 (2.6) | 10 (3.1) | 20 (3.5) |  |  |  |  |  |  |
| **Bocaviruses** | 44 (10.2) | 47 (14.7) | 93 (16.1) | 1.18<br>(0.73, 1.91) | 0.574 | 1.36<br>(0.89, 2.08) | 0.212 | 1.12<br>(0.74, 1.68) | 0.684 |
| **Metapneumo-viruses** | 8 (1.9) | 24 (7.5) | 71 (12.3) | 3.83<br>(1.72, 8.52) | **0.004** | 5.75<br>(3.14, 10.53) | **<0.001** | 1.86<br>(1.17, 2.98) | 0.037 |
| **Rhinoviruses/**<br>**Enteroviruses** | 176 (40.9) | 138 (43.3) | 248 (43.0) | 1.02<br>(0.73, 1.43) | 0.890 | 1.15<br>(0.87, 1.54) | 0.379 | 1.06<br>(0.79, 1.43) | 0.684 |
| ED-68 | 2 (0.5) | 5 (1.6) | 3 (0.5) |  |  |  |  |  |  |
| **Influenzas** | 4 (0.9) | 3 (0.9) | 15 (2.6) |  |  |  |  |  |  |
| IVA | 3 (0.7) | 2 (0.6) | 11 (1.9) |  |  |  |  |  |  |
| IVA-H1 | 0 (0.0) | 0 (0.0) | 1 (0.2) |  |  |  |  |  |  |
| IVA-H1N1 | 1 (0.2) | 2 (0.6) | 4 (0.7) |  |  |  |  |  |  |
| IVA-H3 | 2 (0.5) | 1 (0.3) | 7 (1.2) |  |  |  |  |  |  |
| IVB | 1 (0.2) | 0 (0.0) | 2 (0.3) |  |  |  |  |  |  |
| **Pneumonias** | 1 (0.2) | 0 (0.0) | 0 (0.0) |  |  |  |  |  |  |
| *C. pneumoniae* | 430<br>(100.0) | 319 (100.0) | 577<br>(100.0) |  |  |  |  |  |  |
| *M. pneumoniae* | 1 (0.2) | 0 (0.0) | 0 (0.0) |  |  |  |  |  |  |
| **Parainfluenzas** | 31 (7.2) | 29 (9.1) | 69 (12.0) | 1.42<br>(0.81, 2.47) | 0.440 | 1.88<br>(1.17, 3.03) | **0.015** | 1.61<br>(0.98, 2.65) | 0.118 |
| PIV-1 | 2 (0.5) | 2 (0.6) | 12 (2.1) |  |  |  |  |  |  |
| PIV-2 | 0 (0.0) | 2 (0.6) | 2 (0.3) |  |  |  |  |  |  |
| PIV-3 | 28 (6.5) | 20 (6.3) | 47 (8.1) |  |  |  |  |  |  |
| PIV-4A | 2 (0.5) | 7 (2.2) | 10 (1.7) |  |  |  |  |  |  |
| PIV-4B | 5 (1.2) | 4 (1.3) | 12 (2.1) |  |  |  |  |  |  |
| **RSVs** | 15 (3.5) | 34 (10.7) | 135 (23.4) | 3.17<br>(1.71, 5.87) | **0.002** | 9.28<br>(5.43, 15.85) | **<0.001** | 2.99<br>(1.99, 4.48) | **<0.001** |
| RSV-A | 6 (1.4) | 21 (6.6) | 83 (14.4) |  |  |  |  |  |  |
| RSV-B | 9 (2.1) | 14 (4.4) | 55 (9.5) |  |  |  |  |  |  |
| **SARS-CoV-2** | 38 (8.8) | 19 (6.0) | 15 (2.6) | 0.78<br>(0.42, 1.43) | 0.556 | 0.33<br>(0.16, 0.68) | **0.006** | 0.50<br>(0.25, 0.98) | 0.118 |

In contrast, RSV was detected in 10.7% of mLRIs and 23.4% of sLRIs but occurred in only 3.5% of URIs. Thus, RSV was associated with 3.17-and 9.28-fold increased odds of mLRIs and sLRIs (FDR: 0.002 and <0.001), respectively. RSV also significantly increased the odds of sLRI vs. mLRI (aOR, 2.99, FDR<0.001).

#### High LRI risk viral species

HMPV detections were observed in 7.5% and 12.3% of mLRIs and sLRIs versus only 1.9% of URIs, and thus were associated with 3.83-and 5.75-fold increased odds of mLRI and sLRIs, respectively (FDR=0.004; FDR<0.001). PIV detections were more common in sLRIs (12.0%) than either mLRIs (9.1%) or URIs (7.2%). PIV was associated with 1.88-and 1.61-fold increased risk of sLRI compared to URI and mLRIs, respectively (FDR, 0.015; FDR, 0.118). Non-SARS-CoV-2 coronavirus infections were observed at higher rates in mLRIs (6.9%) and sLRIs (7.8%) than URIs (3.3%) and significantly increased the odds of sLRI vs. URI (sLRI vs URI: aOR, 2.21; FDR=0.015).

#### Low LRI risk viral species

HBoV was detected in a large percentage of both mLRIs (14.7%) and sLRIs (16.1%). However, these frequencies were not significantly higher than URIs (10.2%). SARS-CoV-2 was the only viral species more commonly detected among URIs (8.8%) vs. mLRIs (6.0%) and sLRIs (2.6%). Thus, SARS-CoV-2 infections were associated with 0.33-times lower odds of sLRI (FDR, 0.006). Adenoviruses were detected in 2.3%, 4.1%, and 3.8% of URIs, mLRIs, and sLRIs, respectively, and were not significantly associated with illness severity. Influenza detections were observed in less than 3% of both LRI and URI illnesses and thus were not tested for association with LRIs.

### Viral species co-infections increase severe LRI risk

The prior analyses considered the independent contribution of each viral species to LRI risk. However, co-infection with two or more viral species groups occurred in 22.5% of respiratory illnesses. To examine the influence of viral co-infections on LRI risk, we stratified illnesses into those negative for all viruses, positive for a single virus, and positive for multiple viruses. While infection with a single virus increased the odds of sLRI vs. URI by 1.26-fold (95% CI, 0.92 to 1.74; P=0.16), infection with multiple viruses resulted in 3.68-fold higher odds of sLRI (95% CI, 2.45 to 5.54; P<0.001) relative to illnesses with no detectable viral pathogen (i.e., virus-negative illnesses), and 2.92-fold higher odds (95% CI, 2.05 to 4.16; P<0.001) relative to illnesses positive for a single virus (Figure 3A). Similarly, infection with multiple viruses increased the odds of sLRI vs. mLRI by 1.90-fold (95% CI, 1.26 to 2.87; P=0.002) relative to virus-negative illnesses, and 1.55-fold (95% CI, 1.11 to 2.15; P=0.009) relative to illnesses positive for a single virus.

Examining the characteristics of these co-infections, 78.4% involved 2 viral species, 16.7% involved three, and 4.9% involved four or more viruses. The viruses most common in these co-infections were HRV/EV (74.5%), HBoV (44.4%) and RSV (26.1%). The four most common pairs of viruses all involved HRV/EV, with HRV/EV-HBoV present in 32.4% of co-infections, HRV/EV-RSV present in 15.7%, HRV/EV-PIV in 15.0%, and HRV/EV-HMPV in 13.4%.

### Rhinovirus and Bocavirus co-infections increase Severe LRI risk

We next investigated whether these specific co-infections were associated with increased risk of sLRI vs. URI, and if these viral combinations resulted in higher risk than that conferred by isolated infections with the viral species involved in these co-infections (Figure 3B, eTable 3). For the most common viral combination, isolated infections with HRV/EV or HBoV did not significantly impact sLRI risk (HRV/EV, aOR, 1.04; 95% CI, 0.75 to 1.43; HBoV, aOR, 0.85; 95% CI, 0.48 to 1.50), but these two viruses in combination increased the odds of sLRI by 2.21-fold (95% CI, 1.20 to 4.09, interaction P=0.03). In contrast, isolated RSV infections were associated with higher risk of sLRI (aOR, 11.71; 95% CI, 5.72 to 21.83) compared to when observed in combination with RV/EV (aOR, 5.33; 95% CI, 2.24 to 12.67; interaction P=0.16). SLRI risk was slightly higher for PIV infections observed in combination with HRV/EV (aOR, 2.42; 95% CI, 0.97 to 6.03), as compared to isolated PIV infections (aOR, 1.74; 95% CI, 0.99 to 3.06, interaction P=0.58). Co-infections of these viral species did not significantly alter odds of mLRI vs URI compared to single infections (eFigure 4, eTable 3).

## DISCUSSION

Evaluating respiratory viral pathogens in over 1,300 early-life illnesses, we identified the frequency and risk of LRIs associated with common viral species. A diversity of viruses were detected in LRIs, with several species distinctly associated with sLRIs, including RSV, PIV, HCoV, and HMPV. Though HRV and HBoV were detected in half of sLRIs, these two viruses did not independently increase sLRI risk and were frequently detected in both mLRIs and URIs as well. However, HRV and HBoV co-infections were a significant risk factor for sLRI, as were co-infections more broadly. These differences in risk profiles associated with viral pathogens, in both single and co-infections, suggest multiple mechanisms underlie the development of sLRIs. Moreover, recognizing and monitoring such co-infections could be essential in clinical risk stratification and management.

Respiratory illness surveillance in PRIMERO focused on identifying viral species that differentiate URIs from both mild and severe LRIs. This illness classification system was used as severe LRIs have been strongly associated with asthma development.^1–6^ Our findings confirm that viral infections drive most early-life RIs and are significant risk factors for sLRIs, with certain pathogens contributing disproportionately. RSV was associated with a 3.2-and 9.3-fold increase in the odds of mild and sLRIs, respectively, consistent with cross-sectional studies of bronchiolitis hospitalizations.^23^ These findings underscore the clinical importance of targeted prevention strategies, such as RSV immunoprophylaxis, particularly for high-risk populations. HMPV also conferred a nearly sixfold increased sLRI risk, while common CoV and PIV infections doubled sLRI risk. These findings align with the PREVAIL^24^ and ORChID^25^ early-life cohorts, which reported higher rates of symptomatic vs. asymptomatic infections with RSV, PIV, and HMPV. Together, this evidence highlights the intrinsic pathogenicity of RSV, HMPV, PIV, and to a lesser extent, CoVs in causing sLRIs in infants.

We found HRV infections were common across all illness types and the most frequently detected pathogen in sLRIs, despite not increasing sLRI risk. A previous examination of early-life wheezing illnesses similarly found that HRV was the most commonly detected virus among respiratory illnesses of all severities during the first year of life.^26^ HRV was also the most common detection in both LRIs and asymptomatic infections in the ORCHID study,^25^ while the PREVAIL study found that HRV infections were associated with higher odds of asymptomatic vs. symptomatic infections.^24^ These findings suggest the propensity for HRV to cause severe lower respiratory symptoms is dependent on other host and/or environmental factors. Supporting this, genetic variants at the 17q21 locus influencing ORMDL3 expression are linked to increased early-life HRV-related wheezing risk.^15^ Additionally, a maternal history of atopy has been associated with a 2.4-fold higher risk of HRV bronchiolitis in a cohort of healthy infants, implicating allergic or type-2 biology in more severe HRV pathobiology.^27^ Further studies are needed to directly explore airway/immune mechanisms underlying severe HRV illnesses. Another factor that may influence LRI risk is the subtype of HRV. For example, a study of acute RIs in children under the age of 5, found HRV-C infections were associated with LRIs as compared to HRV-A.^16^ Here, we failed to see evidence of increased severity with HRV-C, perhaps due to the younger age of our cohort.

Similar to HRV, HBoV infections were common among all illness types, including sLRIs, but were not associated with increased sLRI risk. This result supports the notion that HBoV can be pathogenic, disputing original thoughts that it was a “harmless passenger” virus.^28^ A recent Chinese cohort of children hospitalized for either HMPV or HBoV single infections, found HBoV infections were significantly more likely to need mechanical ventilation or ICU admission.^29^ In contrast, other recent large cohort studies have found that HBoV infections are more likely to be asymptomatic than symptomatic.^24, 25^

Multiple viruses were detected in one third of sLRIs in PRIMERO. These co-infections increased the odds of sLRI by 3-fold compared with single virus infections. The ORChID study also found that coinfections increased the odds of LRI.^25^ Interestingly, we found that despite neither HBoV nor HRV single infections being associated with sLRI risk, co-infections between these pathogens increased odds of sLRI by over 2-fold. Supporting this, in the ORChID cohort, viral co-infections largely involved the combination of HRV and a DNA virus, with rhinovirus/bocavirus co-infection the most common among symptomatic infections. Additionally, an Italian study of 165 children hospitalized with HBoV mono-infection or co-infection, found those with co-infection were at higher risk of ICU admission and experienced a longer length of stay.^30^ Together, these findings reinforce the importance of viral surveillance in identifying high-risk co-infections and guiding decisions around hospitalization or intensive monitoring. Further studies are needed to understand the basis of increased severity for viral co-infections, which could involve: (1) one virus altering the host immune response, allowing a more exuberant infection by the subsequent virus, (2) additive effects of multiple distinct pathobiologic mechanisms for two viruses, or (3) deleterious synergy, either in the form of viral pathogenic mechanisms or host responses.

PRIMERO’s surveillance period, spanning the start of the pandemic through the end of public health mitigation measures, allowed us to assess the impact of reduced viral circulation on LRI rates in infancy. Among children born during the first year of the pandemic, all respiratory illness rates declined, with LRIs significantly reduced compared to children born after the first year of the pandemic. This likely reflects the decrease in circulation of high-risk viruses like RSV and HMPV during this period, while low-risk species like HRV and SARS-CoV-2 showed continued, though limited circulation. Given the established link between early-life sLRIs and childhood asthma,^1–6^ follow-up studies in PRIMERO and similar cohorts could reveal whether infants born during pandemic restrictions exhibit reduced asthma rates.

Surveilling RIs through the COVID pandemic also allowed us to find that SARS-CoV-2, uniquely among viral species, was more common in URIs, suggesting the infant immune response to SARS-CoV-2 is particularly suited to control these infections. Supporting this, we and others have found that SARS-CoV-2 infections are much more likely to be asymptomatic in infants than in adults.^19^ Although the reason for these results is unclear, one possible explanation is the increased interferon and innate immune responses among infants, to which SARS-CoV-2 has been shown to be particularly sensitive.^31, 32^

This study has several limitations. The PRIMERO cohort was recruited from a single hospital in Puerto Rico; therefore, rates of early-life respiratory illnesses, frequencies of viral detections, and seasonal patterns of illnesses and viruses, may not be generalizable to other geographic locations. Early-life respiratory illnesses are notably more severe among Puerto Rican children, who face higher rates of asthma and respiratory morbidity due to a combination of biological, environmental, and social factors, including disparities in healthcare access, socioeconomic status, and environmental exposures.^20, 21, 33–35^ Like other cohort studies, we note our reliance on parent report of symptoms to classify illnesses into LRIs and URIs, which may have led to misclassification, particularly between URIs and mLRIs. Our viral screen of illnesses was limited to a subset of illnesses with available swabs from in person assessments. Additionally, our viral detection rates may have been influenced by alterations in patterns of viral circulation and respiratory illnesses during the early part of the PRIMERO study, which was conducted during COVID lockdowns.

### Conclusions

Early-life sLRIs are caused by diverse viral pathogens. RSV, HMPV, PIV, and common CoV species are associated with increased sLRI risk, while SARS-CoV-2 infections are associated with decreased risk. Though HRV and HBoV infections trigger the majority of sLRIs, these species are not by themselves associated with increased risk; therefore, their pathogenicity may be dependent on other host and environmental factors. Viral co-infections, particularly those involving HRV and HBoV, are associated with elevated sLRI risk. These results underscore the heterogeneity of viral species driving development of sLRIs in infants and underscore the need for additional investigation of the biological bases underlying this heterogeneity. Prioritizing early detection of high-risk pathogens and co-infections can guide timely interventions to mitigate severe outcomes in vulnerable infants.

## Supporting information

eTable, eFigure, eMethods

## Data Availability

All data produced in the present study are available upon reasonable request to the authors, via the PRIMERO Data Coordinating Center at University of California San Francisco.

## Acknowledgement

Author Contributions: Drs. Moore and Seibold had full access to all of the data in the study and take responsibility for the integrity of the data and the accuracy of the data analysis.

## Concept and design

Seibold, Moore, Sheppard, Rodriguez-Santana

*Acquisition, analysis, or interpretation of data:* Moore, Seibold, Secor, Fairbanks-Mahnke, Everman, Elhawary, Witonsky, Pruesse, Jackson, Li, Morin, Sajuthi, Williams, Sheppard, Rodriguez-Santana, Chang, Contreras, Eng, Canales, Rosado, Hu, Scott Huntsman, Lopez, Valentin, Medina, Angely Montanez-Lopez, Morin, Nieves, Oh, Otero, Colon, Rodriguez, Salazar, Serrano, EV Morales, Gabriela Vazquez, NV Morales, Sheppard, Rodriguez-Santana

## Drafting of the manuscript

Moore, Seibold, Secor

## Critical review of the manuscript for important intellectual content

All authors

## Statistical analysis

Moore, Secor

## Obtained funding

Seibold, Rodriguez Santana *Administrative, technical, or material support:* Elhawary, Eng *Supervision:* Seibold, Moore, Sheppard, Rodriguez-Santana

## Additional Contributions

The authors acknowledge the families for their participation and thank the numerous data collectors, recruiters, technicians, and hospital administrators for their support and participation in PRIMERO.

