## Supplementary material for "Independent and interactive effects of viral species on risk for lower respiratory tract illnesses in early life": eTable, eFigure, eMethods

***Study design***

Healthy mother-infant pairs were enrolled from the Hospital Interamericano de Medicina Avanzada-San Pablo (HIMA) in Caguas, Puerto Rico, which neighbors the capital of San Juan. Pregnant women aged 18 years or older, residing in Puerto Rico, and planning to deliver at HIMA were invited to participate in the study and provided written consent. To be eligible, newborns had to live with their biological mother, be afebrile (<38°C), without clinical signs of respiratory infection, at least 37 weeks of gestational age, and have a birthweight of at least 2,500 g. The PRIMERO protocol and informed consent documents were approved by the UCSF Institutional Review Board (IRB; #18-26263) and by a National Institutes of Health (NIH)-appointed independent Observational Study Monitoring Board (OSMB).

***Illness Surveillance***

Mothers received a weekly automated SMS text or email with a hyperlink to indicate whether the child has signs of respiratory illness. Mothers reporting that their child had signs of illness were contacted by project staff with a phone call to administer a screening questionnaire to capture LRIs based on the presence of symptoms, including a cough that interferes with daily activities, wheezing/whistling in the chest, fast breathing or gasping for air, and sleep disturbed by cough, wheeze, or difficulty breathing. Participants reporting only a runny/plugged nose, sneezing, and/or a mild cough were considered most likely to have an upper respiratory tract illness (URI). All LRIs, as well as an infant's first URI each year, were invited for an in-person clinic assessment. During in-person visits, a nasal swab was collected and symptoms were assessed.

All illnesses were tracked until resolution by weekly telephone questionnaire in order to determine the trajectory of the illness, the continued presence and severity of symptoms, and the need for hospitalization or systemic steroid treatment. A final illness classification of LRI vs. URI was made for each illness event based on the symptoms reported across all assessments conducted for the illness. LRIs reported one or more of the following symptoms during the course of the illness: cough that interferes with daily activities, wheezing/whistling in the chest, fast breathing or gasping for air, and/or sleep disturbed by cough, wheeze, or difficulty breathing. LRIs were considered severe if the child required hospitalization or treatment with systemic corticosteroids for the illness and mild otherwise.

***Nasal swab nucleic acid extraction***

Contoured pediatric flocked swabs with a stopper (Copan diagnostics; 56780CS01) were used to collect airway epithelium from the posterior surface of the inferior turbinate during illness visits for participants under one year in age. Minitip flocked swabs without a stopper (Copan diagnostics; 518CS01) were used to collect airway epithelium from the posterior surface of the inferior turbinate from children for illness events occurring at greater than one year in age. Collected swabs were immediately collected into RLT Plus lysis buffer (Qiagen) supplemented with 40mM Dithiothreitol, vortexed, and frozen at

−80°C until extraction, for complete lysis of sampled cells and microorganisms (host epithelial cells, viral particles, etc.). RNA and DNA were extracted using the Zymo Quick DNA/RNA mini kit extraction protocol adapted for use on the Biomek i7 automated liquid handler system. Extracted nucleic acids were stored at -80°C until testing.

**Respiratory Pathogen Panel (RPP) testing of nasal swabs**

RNA and DNA from illness swabs were used to assay for 21 pathogens using the NxTAG Respiratory Pathogen Panel (RPP; Diasorin). Briefly, 3ul of RNA and 3ul of DNA were combined for each sample and mixed with 2ul of MS2 internal assay control and 29ul of molecular biology grade water of a total volume of 37ul. From each sample mix, 35ul was added to lyophilized reaction beads, amplified by PCR, and data captured using the MagPix system as per manufacturer's instructions. Multiplexed RPP kits used in this study include viral detection of Influenza (A, A H1, A H3, and B), Respiratory Syncytial Virus (A and B), Parainfluenza virus (1, 2, 3, 4A and 4B), Human Bocavirus (HBoV), Human Metapneumovirus (HMPV), Rhinovirus/Enterovirus (HRV/EV), Adenovirus (AdV), and Coronavirus (HKU1, NL63, 229E, and OC43), and bacterial detection of *Chlamydophila pneumoniae* and *Mycoplasma pneumoniae*.

**SARS-CoV-2 and ED68 testing**

Samples were tested for the SARS-CoV-2 infection using the CDC assays for N1 and N2 of SARS-CoV-2. Enterovirus D68 (ED68) was measured using the CDC 2022 ED68 assay (CDC),[1] and multiplexed with the housekeeping control RNaseP. For assay input,

4ul RNA, 4ul DNA, and 3ul water were mixed to generate the qPCR input template. PCR reactions were assembled using the Echo525 acoustic liquid handler (Labcyte) as 2.5ul reaction volumes using 0.25 ul of nucleic acid template mix, 1-step ToughMix low Rox qPCR master mix (QuantaBio), and specific primers and probes for N1 and N2, or ED68 and RnaseP, SARS-CoV-2 N1 and N2 assays were duplexed as previously described,[2] and amplified as follows: 10 minutes at 50°C, 3 minutes at 95°C, followed by 45 cycles of denaturation at 95°C for 3 seconds and annealing for 30s at 58°C. The CDC 2022 ED68 assay and RNAseP assays were duplexed, and amplified as follows: 10 minutes at 50°C, 3 minutes at 95°C, followed by 45 cycles of denaturation at 95°C for 3 seconds and annealing for 30s at 52°C. RT-qPCR results were run on a QuantStudio 7Pro and analyzed using the QuantStudio Design and Analysis software.

**Human Rhinovirus Typing**

Nasal RNA that tested positive for HRV/EV by the RPP assays were selected for human rhinovirus (HRV) subtype determination as previously described.[3] Reactions were assembled and miniaturized using the Biomek i7 automated liquid handler fitted with an Echo525 acoustic liquid handler (Labcyte), and amplified on an Eppendorf Mastercycler X50s. For all HRV/EV positive samples, RNA was diluted 1:5, and 5ul of diluted RNA was transcribed to cDNA in a 10ul reaction volume using the High-capacity cDNA reverse transcription kit with RNase inhibitor (Applied Biosystems, #4374966) as per manufacturer's instructions. PCR reaction 1 was assembled as a 8ul reaction with 0.8ul of cDNA template using the Platinum PCR SuperMix HF (Invitrogen #12532016) using primers HRV-B1, HRV-B2, HRV-B3, and 5UTR-rev (eTable 4) and amplified as follows:

94°C for 2min, followed by 16 cycles of 94°C denaturation for 20sec, touchdown annealing for 30sec from 68-54°C decreasing the temperature by 2°C every 2 cycles, and extension at 68°C for 40sec, followed by 11 cycles of denaturation at 94°C for 20sec, anneal for 30sec at 52°C, and extension at 68°C for 40 sec. The nested PCR reaction 2 was assembled as a 5ul reaction using 0.5ul of template from PCR 1 with GoTaq Colorless Master Mix (Promega #M7132), primers 5UTRn-A1, 5UTRn-A2, 5UTR-nA3, 5UTRn-B1, 5UTRn-B2, 5UTRn-Cc, and 5UTRn-rev (eTable 4), and amplified using the cycling conditions as follows: 94°C for 2 min, followed by 32 cycles of 94°C denaturation for 20 sec, annealing at 52°C for 30 sec, and extension at 68°C for 40 sec, with a final extension at 3 min at 68°C. PCR 2 product cleanup was conducted using ExoSAP-IT reagent (Applied Biosystems #78200.200.UL) as per the manufacturer's instructions. HRV amplification positivity of the 375bp product was determined using a 1.5% agarose gel, and the product was directly sequenced using the 5UTR-rev primer (McLab, San Francisco, CA).

Basecalling on Sanger chromatogram trace files was performed with Tracy[4] (version 0.7.6, trimming stringency 7, retrieving the primary sequence only). The resulting sequences were aligned using SINA[5] (version 1.7.2) and phylogenetically placed and classified with pplacer, guppy and rppr[6] (version v1.1.alpha19, MRCA mode).

HRV subtype was confidently determined if the SINA normalized alignment score for the sample was 70 or greater, the sequence had a length between 250 and 500bp, had an N count of 10 or fewer, and had a species posterior probability of 0.6 or greater.

We employed a custom classification reference comprising 643 taxa covering HRV-A, B, and C, as well as 3 outgroup taxa from Enterovirus A, based on the Palmenberg HRV phylogeny.[7] The initial multiple sequence alignment construction was performed with MaFFT, the phylogenetic tree was inferred with RAxML[8] (version 8.2), alignment curation and masking as well as taxonomic annotation of the phylogenetic tree were performed with ARB[9] version 7.1 and the pplacer classification reference package constructed with taxstastic.[6]

***Statistical analysis***

Participant demographics, event characteristics, and viral detections were summarized by frequency and percent or median and interquartile range (IQR). The probability of experiencing one or more URIs, mild LRIs, and severe LRIs over the first two years of life were calculated using Kaplan-Meier methods, separately for children born during the first year of the pandemic (before March 1, 2021) and after the first year and these groups were compared using the log-rank test. We tested for association between odds of severe LRI vs. URI, mild LRI vs. URI, and severe LRI vs. mild LRI and virus species using generalized estimating equation (GEE) logistic regression models with an exchangeable working correlation structure grouped by participant, as participants could have more than one illness included in the analysis. These models adjusted for age, sex, race and time from illness onset to nasal swab and included indicator variables for the following viral detections: adenoviruses, coronaviruses other than SARS-CoV-2 (229E, HKU1, NL63, or OC43), bocaviruses, metapneumoviruses, rhinoviruses/enteroviruses, parainfluenzas (1,

2, 3, 4A, or 4B), respiratory syncytial viruses (A or B), and SARS-CoV-2. To account for the multiple statistical tests performed, p-values for the 8 virus types were adjusted using a Benjamini-Hochberg correction to control the false discovery rate. We tested for effect modification between viruses in top three observed coinfections (coinfections detected in >3% of illnesses) by adding interaction terms to the model described above. Similar models compared the odds of mild and severe LRI vs. URI between illnesses with no, single, and multiple viral detections. All analyses were performed in R (r-project.org, version 4.2.2), using the geepack,[10] survival,[11] and epiR[12] packages.

**eFigure 1.** Proportion experiencing no, 1, or 2 or more mild and severe LRIs among children reaching 2 years of age. Children that experienced at least one mild (or severe) LRI, but that had additional LRIs with missing severity data were classified as having an "Unknown Number" of mild (or severe) LRIs. In addition, it could not be determined if some children experienced either type of LRI due to missing symptom or severity data for one or more illnesses; these participants were classified as "Unknown Status".

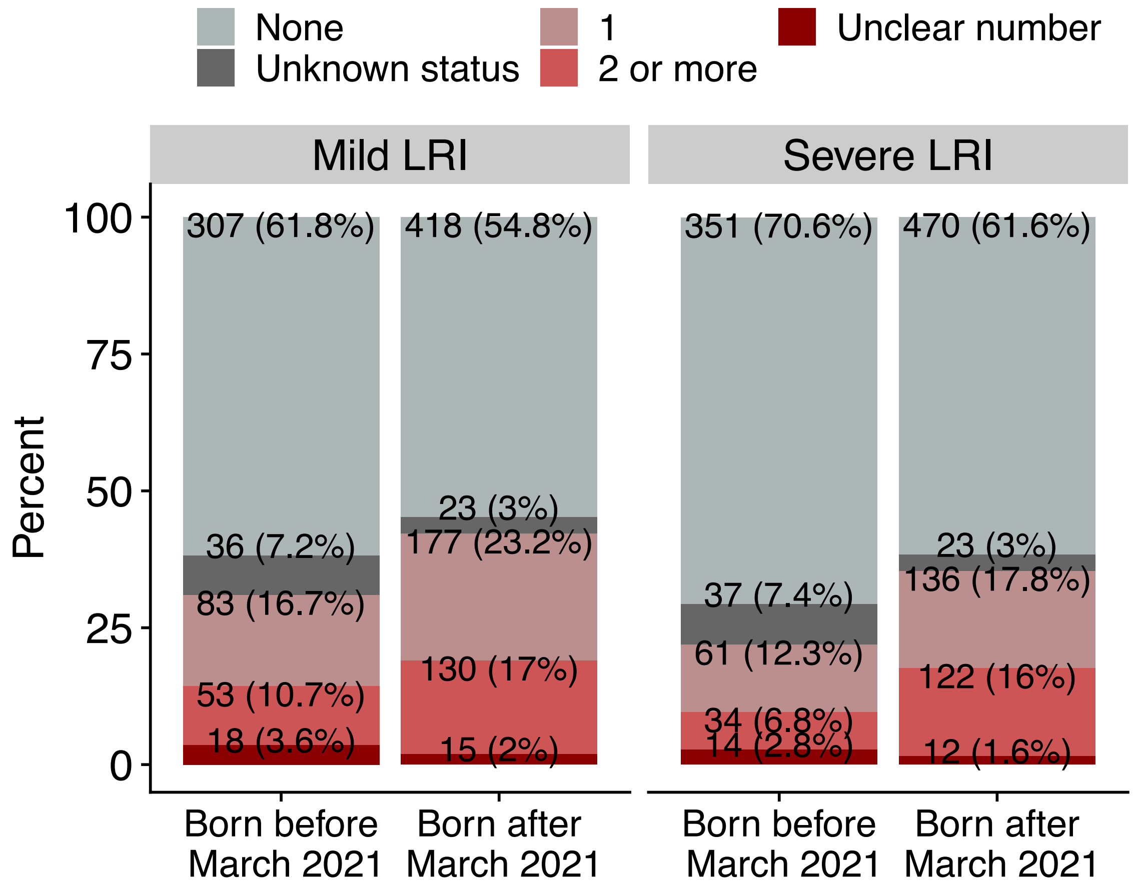

**eFigure 2.** Overview of samples included in viral determinants of LRI analyses.

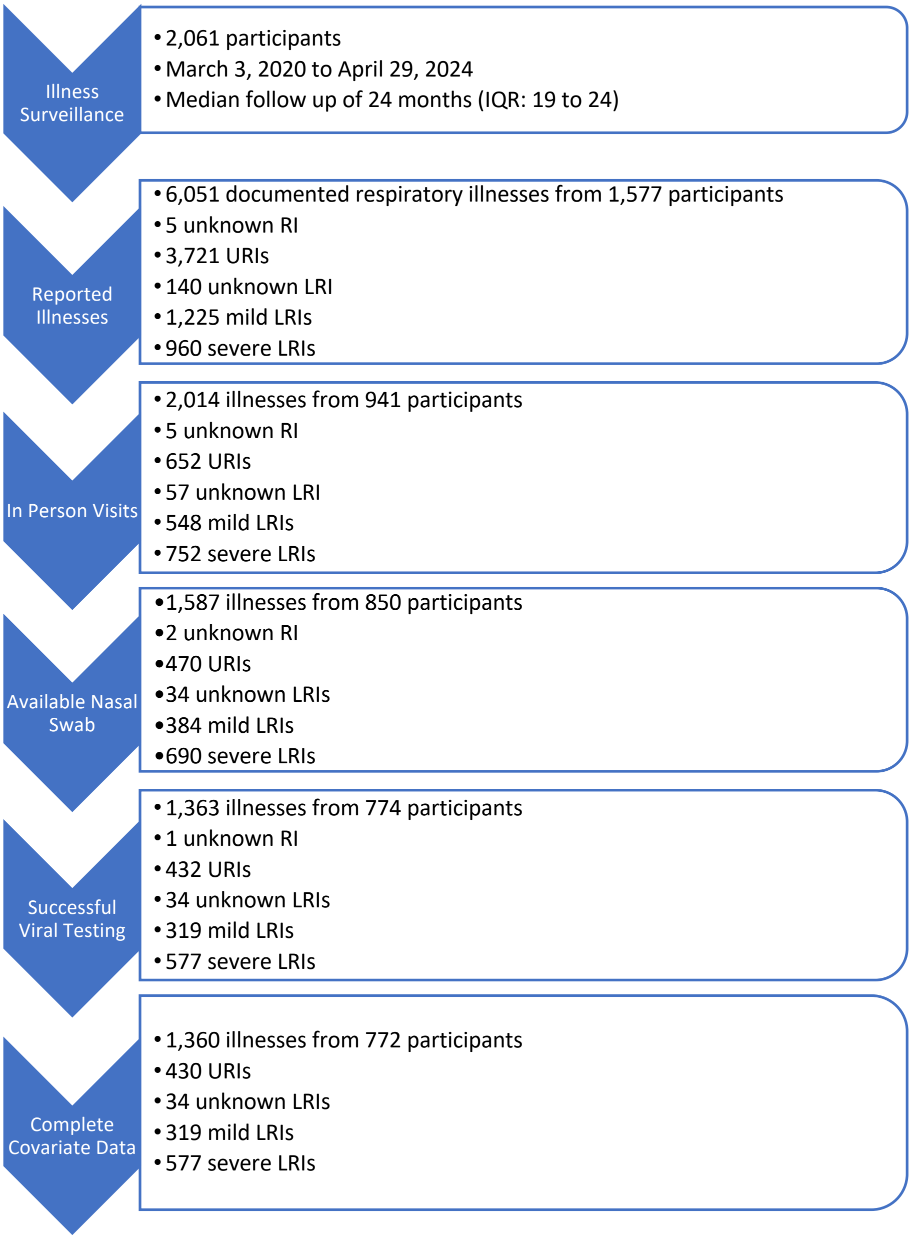

**eFigure 3.** Rhinovirus typing by illness type. N (%).

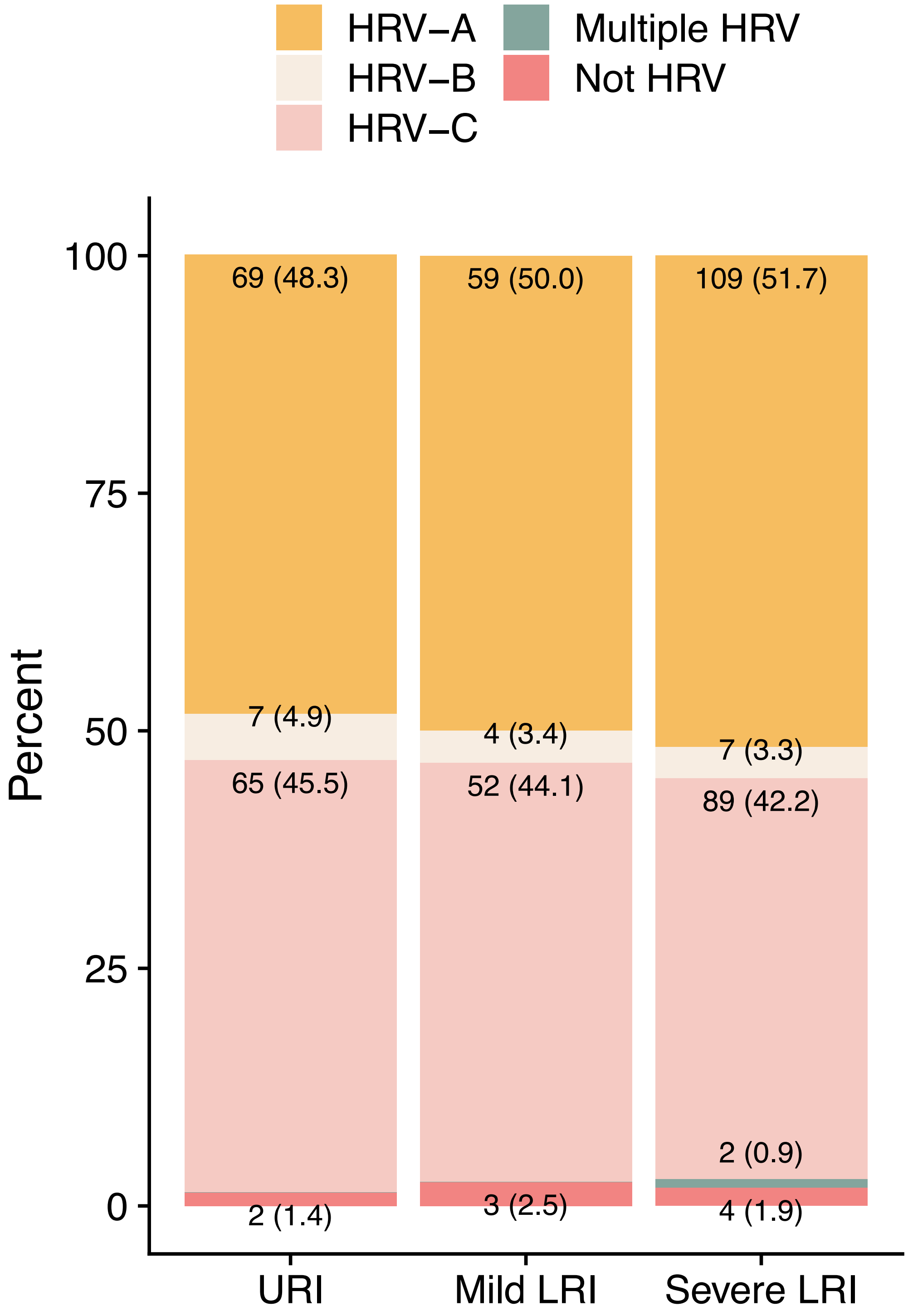

**eFigure 4.** Forest plot of adjusted odds ratios comparing the odds of mild LRI vs URI, severe LRI vs. URI, and severe LRI vs. mild LRI between illnesses that test positive and negative for specific viral infections and co-infections. Blue dots represent the estimated aOR and whiskers indicate the 95% confidence interval.

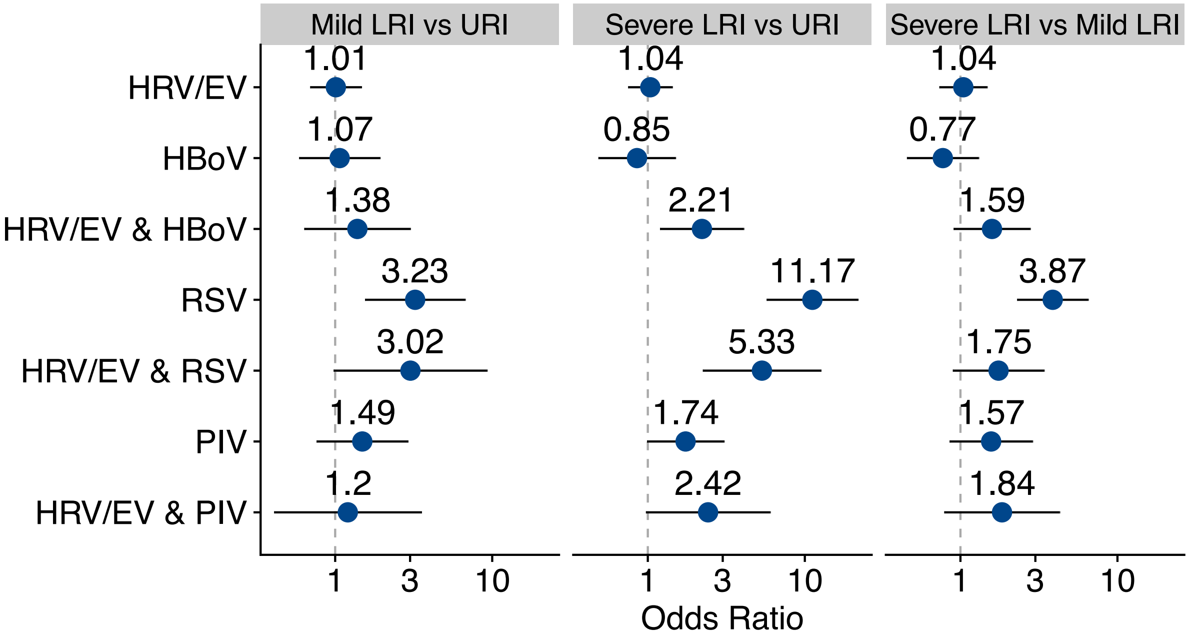

**eTable 1.** Characteristics of illness events with viral testing results in the PRIMERO cohort from March 3, 2020 to April 29, 2024. N (%) or median [interquartile range].

| | URI (N=430) | Mild LRI (N=319) | Severe LRI (N=577) | LRI of Unknown Severity (N=34) |
|---|---|---|---|---|
| **Virus Positive** | 286 (66.5) | 240 (75.2) | 468 (81.1) | 14 (41.2) |
| **Male** | 230 (53.5) | 158 (49.5) | 350 (60.7) | 18 (52.9) |
| **Race** | | | | |
| *Black* | 103 (24.0) | 86 (27.0) | 141 (24.4) | 11 (32.4) |
| *Other/Mixed* | 173 (40.2) | 129 (40.4) | 222 (38.5) | 14 (41.2) |
| *White* | 154 (35.8) | 104 (32.6) | 214 (37.1) | 9 (26.5) |
| **Age at Illness Onset** (Months) | 8.00 [4.00, 13.00] | 10.68 (6.11) | 11.00 [7.00, 16.00] | 6.00 [2.00, 12.50] |
| **Time from Illness Onset to Nasal Swab** (Days) | 12.00 [7.00, 19.00] | 11.70 (8.54) | 10.00 [5.00, 17.00] | 12.00 [9.00, 19.75] |
| **Tobacco Smoke Exposure in Household** | 60 (14.0) | 44 (13.8) | 61 (10.6) | 8 (23.5) |
| **Parental History of Atopy** | 313 (73.0) | 243 (76.2) | 428 (74.3) | 28 (84.8) |
| **Season** | | | | |
| *Summer* | 112 (26.0) | 68 (21.3) | 99 (17.2) | 5 (14.7) |
| *Fall* | 118 (27.4) | 103 (32.3) | 266 (46.1) | 14 (41.2) |
| *Winter* | 94 (21.9) | 79 (24.8) | 113 (19.6) | 8 (23.5) |
| *Spring* | 106 (24.7) | 69 (21.6) | 99 (17.2) | 7 (20.6) |
| **First Year of Pandemic** | 20 (4.7) | 5 (1.6) | 6 (1.0) | 6 (17.6) |
| **Hospitalization** | 21 (11.3) | 0 (0.0) | 175 (30.5) | |
| **Systemic Steroids** | 36 (19.6) | 0 (0.0) | 508 (89.4) | |
| **Symptoms** | | | | |
| *Runny Nose* | 393 (91.4) | 301 (94.4) | 553 (95.8) | 29 (85.3) |
| *Sneezing* | 175 (40.7) | 196 (61.4) | 406 (70.4) | 7 (20.6) |
| *Cough* | 329 (76.5) | 302 (94.7) | 573 (99.3) | 26 (76.5) |
| *Cough interfering with daily activities* | 0 (0.0) | 151 (47.3) | 435 (75.4) | 8 (23.5) |
| *Wheezing* | 0 (0.0) | 109 (34.2) | 439 (76.1) | 2 (5.9) |
| *Fast breathing or gasping for air* | 0 (0.0) | 139 (43.6) | 424 (73.5) | 13 (38.2) |
| *Sleep disturbed by cough, wheeze or difficulty breathing* | 0 (0.0) | 247 (77.4) | 495 (85.8) | 19 (55.9) |

**eTable 2.** Comparison of odds of viral species detection between seasons among respiratory illnesses screened after February 28, 2021, adjusting for sex, age, race, and time from illness onset to nasal swab.

| **Virus** | **F-test for Difference Between Seasons** | | **Fall vs. Summer** | | **Winter vs Summer** | | **Spring vs. Summer** | | **Fall vs. Winter** | | **Fall vs Spring** | | **Winter vs Spring** | |
|---|---|---|---|---|---|---|---|---|---|---|---|---|---|---|
| | **P-Value** | **FDR** | **OR (95% CI)** | **FDR** | **OR (95% CI)** | **FDR** | **OR (95% CI)** | **FDR** | **OR (95% CI)** | **FDR** | **OR (95% CI)** | **FDR** | **OR (95% CI)** | **FDR** |
| HRV/EV | 0.697 | 0.697 | 0.95 (0.7, 1.28) | 0.793 | 1.13 (0.8, 1.6) | 0.793 | 1.05 (0.75, 1.47) | 0.793 | 0.84 (0.62, 1.14) | 0.793 | 0.91 (0.68, 1.21) | 0.793 | 1.08 (0.76, 1.53) | 0.793 |
| RSV | **<0.001** | **<0.001** | 6.21 (3.61, 10.7) | **<0.001** | 1.41 (0.73, 2.72) | 0.310 | 0.45 (0.19, 1.06) | 0.080 | 4.42 (2.78, 7.02) | **<0.001** | 13.87 (6.63, 29.04) | **<0.001** | 3.14 (1.38, 7.12) | **0.009** |
| PIV | 0.101 | 0.188 | 0.82 (0.48, 1.38) | 0.545 | 1.12 (0.63, 1.99) | 0.698 | 1.51 (0.87, 2.61) | 0.394 | 0.73 (0.44, 1.22) | 0.394 | 0.54 (0.33, 0.88) | 0.081 | 0.74 (0.44, 1.25) | 0.394 |
| HBoV | **<0.001** | **<0.001** | 2.21 (1.16, 4.2) | **0.023** | 8.11 (4.28, 15.35) | **<0.001** | 1.46 (0.71, 3.01) | 0.302 | 0.27 (0.18, 0.4) | **<0.001** | 1.51 (0.89, 2.55) | 0.147 | 5.54 (3.38, 9.1) | **<0.001** |
| HMPV | 0.241 | 0.31 | 1.63 (0.9, 2.95) | 0.315 | 1.01 (0.5, 2.03) | 0.978 | 1.32 (0.69, 2.53) | 0.537 | 1.62 (0.92, 2.85) | 0.315 | 1.23 (0.72, 2.11) | 0.537 | 0.76 (0.39, 1.47) | 0.537 |
| CoV | **<0.001** | **<0.001** | 0.55 (0.24, 1.25) | 0.183 | 2.46 (1.22, 4.97) | **0.024** | 2.30 (1.14, 4.63) | **0.029** | 0.22 (0.11, 0.44) | **<0.001** | 0.24 (0.12, 0.46) | **<0.001** | 1.07 (0.62, 1.85) | 0.805 |
| Inf | 0.272 | 0.31 | 2.47 (0.54, 11.34) | 0.365 | 0.79 (0.11, 5.57) | 0.817 | 2.97 (0.61, 14.49) | 0.357 | 3.11 (0.7, 13.76) | 0.357 | 0.83 (0.34, 2.07) | 0.817 | 0.27 (0.05, 1.32) | 0.357 |
| SARS-CoV-2 | 0.118 | 0.188 | 0.48 (0.25, 0.91) | 0.154 | 0.92 (0.47, 1.81) | 0.905 | 0.88 (0.45, 1.71) | 0.905 | 0.52 (0.26, 1.04) | 0.159 | 0.54 (0.27, 1.07) | 0.159 | 1.04 (0.52, 2.08) | 0.905 |

**eTable 3.** Frequencies of specific single infections and co-infections among URIs, mild LRIs, and severe LRIs and odds ratios comparing the odds of LRI between illnesses that test positive for single viruses and coinfections relative to illnesses that test negative for these viruses. Odds ratios are adjusted for age at illness onset, sex, race, and time from illness onset to nasal swab. P-values for interaction terms are shown in parentheses.

| **Infection** | **URI N (%)** | **MLRI N (%)** | **SLRI N (%)** | **SLRI vs URI** | | **MLRI vs URI** | | **SLRI vs MLRI** | |
|---|---|---|---|---|---|---|---|---|---|
| | | | | **aOR (95% CI)** | **P** | **aOR (95% CI)** | **P** | **aOR (95% CI)** | **P** |
| HRV/EV only | 144 (33.5) | 79 (24.8) | 110 (19.5) | 1.04 (0.75, 1.43) | 0.8253 | 1.01 (0.69, 1.47) | 0.9573 | 1.04 (0.73, 1.48) | 0.8133 |
| RSV only | 9 (2.1) | 20 (6.3) | 75 (13.0) | 11.17 (5.72, 21.83) | **<0.0001** | 3.23 (1.55, 6.74) | **0.0018** | 3.87 (2.3, 6.52) | **<0.0001** |
| HRV/EV & RSV | 4 (0.9) | 12 (3.8) | 32 (5.5) | 5.33 (2.24, 12.67) | **0.0001** (0.1626) | 3.02 (0.98, 9.3) | 0.0547 (0.9060) | 1.75 (0.9, 3.42) | 0.1018 (0.0558) |
| PIV only | 20 (4.7) | 17 (5.3) | 23 (4.0) | 1.74 (0.99, 3.06) | 0.0550 | 1.49 (0.76, 2.91) | 0.2446 | 1.57 (0.85, 2.88) | 0.1471 |
| HRV/EV & PIV | 9 (2.1) | 9 (2.8) | 28 (4.9) | 2.42 (0.97, 6.03) | 0.0573 (0.5827) | 1.20 (0.41, 3.55) | 0.7362 (0.7237) | 1.84 (0.79, 4.29) | 0.1579 (0.8158) |
| HBoV only | 22 (5.1) | 15 (4.7) | 14 (2.4) | 0.85 (0.48, 1.5) | 0.5840 | 1.07 (0.59, 1.93) | 0.8284 | 0.77 (0.46, 1.31) | 0.3370 |
| HRV/EV & HBoV | 18 (4.2) | 22 (6.9) | 56 (9.7) | 2.21 (1.2, 4.09) | **0.0113 (0.0257)** | 1.38 (0.64, 3.01) | 0.4123 (0.5940) | 1.59 (0.91, 2.79) | 0.1065 (0.0604) |

**eTable 4.** Primers and Probes for qPCR and PCR Assays

| SARS-CoV-2 and RNaseP qPCR Primers and Probes | | |
|---|---|---|
| Primer/probe name | Sequence | Final Concentration |
| 2019-nCoV_N1-F | GACCCCAAAATCAGCGAAAT | 500 nM |
| 2019-nCoV_N1-R | TCT GGT TAC TGC CAG TTG AAT CTG | 500 nM |
| 2019-nCoV_N1-P | /5YakYel/ACCCCGCAT/ZEN/TACGTTTGGTGGACC/3IABkFQ/ | 125 nM |
| 2019-nCoV_N2-F | TTACAAACATTGGCCGCAAA | 500 nM |
| 2019-nCoV_N2-R | GCGCGACATTCCGAAGAA | 500 nM |
| 2019-nCoV_N2-P | /56-FAM/ACAATTTGC/ZEN/CCCCAGCGCTTCAG/3IABKFQ/ | 125 nM |
| ED68 CDC2022 Sense (AN993) | GGA ATA AAT CCA GCN GAY CAN AT | 500 nM |
| ED68 CDC2022 Rev | CCA CGC TTT TAT RTG YTT NGG YTT CAT | 500 nM |
| ED68 CDC2022 probe (AN992) | /5HEX/GARCAYCAR/ZEN/CCARTTGGTTTCACAGTGAC/3IABkFQ/ | 500nM |
| RNaseP_Forward | AGATTTGGACCTGCGAGCG | 500 nM |
| RNaseP_Reverse | GAGCGGCTGTCTCCACAAGT | 500 nM |
| RNaseP_Probe | /56-FAM/TTCTGACCT/ZEN/GAAGGCTCTGCGCG/3IABKFQ/ | 125 nM |
| HRV Typing Primers | | |
| Primer name | Sequence | Final concentration |
| HRV-B1 | CAAGCACTTCTGTTTCCCC | 480 nM |
| HRV-B2 | CAAGCACTTCTGTTACCCC | 480 nM |
| HRV-B3 | CAAGCACTTCTGTCTCCCC | 480 nM |
| 5UTRn-A1 | GCACTTCTGTTTCCCCGGY | 480 nM |
| 5UTRn-A2 | GCACTTCTGTTACCCCGGT | 480 nM |
| 5UTRn-A3 | GCACTTCTGTTTCCCCCG | 480 nM |
| 5UTRn-B1 | ACTTCTGTTTCCCCGGAGC | 480 nM |
| 5UTRn-B2 | ACTTCTTGTTCCCCGGAGC | 480 nM |
| 5UTRn-Cc | TTCTGTTTCCCCGGRYGTG | 480 nM |
| 5UTR-rev | ACGGACACCCAAAGTAGT | 480 nM |

10. Halekoh U, Hojsgaard S, Yan J. The R Package geepack for Generalized Estimating Equations. J Stat Softw. 2006;15(2):1-11. doi: DOI 10.18637/jss.v015.i02. PubMed PMID: WOS:000235180600001.
11. Therneau T. A Package for Survival Analysis in R. version 3.5-0 ed2023.
12. Stevenson M, Sergeant E, Firestone S. epiR: Tools for the Analysis of Epidemiological Data. R package 2024.
